# Sexual health in women with Parkinson’s disease: Motor, non-motor, and social impacts

**DOI:** 10.1101/2023.08.30.23294846

**Authors:** Kátia Cirilo Costa Nobrega, Isaíra Almeida Pereira da Silva Nascimento, Bruno Rafael Antunes Souza, Raissa Amorim Gonçalves, Thalyta Silva Martins, Geovanna Ferreira Santos, Bruno Eron de Almeida da Silva, André Frazão Helene, Antonio Carlos Roque, Rodolfo Savica, Maria Elisa Pimentel Piemonte

## Abstract

**Background:** Sexual dysfunction (SD) is a common non-motor symptom (NMS) in people with Parkinson’s disease (PwPD). Sexual health (SH) depends on several biological, mental, and social factors that PD may affect. Despite its prevalence and relevance for quality of life, SD in women with Parkinson’s disease (WwPD) is poorly understood, and research in this area is scarce.

**Objectives:** To investigate the impact of motor, non-motor, and social aspects on the SH of WwPD.

**Methods:** We conducted a cross-sectional study of 100 women (mean age 54.45±8.31, mean H&Y stage 1.70±0.71). The following data were collected for each person (used tests/scales indicated within parentheses and defined in Glossary): (1) demographic information and global cognitive capacity (T-MoCA); (2) non-motor aspects of daily life experiences (MDS-UPDRS, part I); (3) motor aspects of daily life experiences (MDS-UPDRS, part II); (4) fatigue (FSS); (5) self-esteem (RSES); (6) sleep disorder (PDSS); (7) couple relationship quality (CRQ) (DAS); (8) depressive signals (BDI); (8) short-term sexual health (FSFI); and (9) long-term sexual health (SQ-F).

**Results:** Our results suggest that depressive symptoms, preserved cognitive status, and CRQ are predictive factors in the sexual health of WwPD. Age, disease onset, duration, postmenopausal, Levodopa dosage, motor disability, and fatigue were not correlated with SH.

**Conclusion:** Our findings emphasize the need to assess the sexual functioning of WwPD to investigate which motor, non-motor, and social aspects may be involved in SD so that measures can be implemented in clinical practice.

---

Parkinson’s disease (PD) is the second most common neurodegenerative disease [1], affecting over 10 million people worldwide [2]. Clinically, PD has been defined by heterogeneous motor symptoms [3]. Besides motor dysfunctions, people with PD (PwPD) experience diverse non-motor symptoms (NMS), including sexual dysfunction (SD) [4,5]. Although poorly understood and investigated [5], SD is reported as one of the 12 most bothersome symptoms out of 24 analyzed [6]. Furthermore, SD contributes significantly to loss of quality of life in PwPD, increasing medical, social, and economic burden for patients, sex partners, and caregivers [7]. The negative SD impact on quality of life has been demonstrated by several studies [8,9].

Sexual health (SH) is a complex phenomenon in which, besides organic causes, environmental and psychological factors may also be involved. However, data on sexual function in PD are incomplete and often contradictory [10].

In the absence of any illness, the aging process plays a key role in the development of SD [11]. Age is an independent predictive factor for developing erectile dysfunction in men and SD in women [12]. However, the aging effect on SH in PD is a complex issue and several studies showed that SD and sexual satisfaction in PD worsened with aging [13,14,15,16]. The PD age of onset appears to influence SH too. People with early-onset PD (EOPD) reported more SD [17,18,19,20,21] and more decrease in libido and increased difficulty in sexual activities [22] than people with late-onset PD (LOPD).

Gender is another confusing factor for SD. While some studies found higher prevalence of SD in men with PD (MwPD) than in women with Parkinson’s disease (WwPD) [8,23], others showed prevalence ranging from 42.6% to 79% in MwPD and 36% to 87.5% in WwPD [14,24,25,26]. In contrast, a study showed WwPD reported higher SD than MwPD [21] while another revealed that overall sexual function in PwPD is similar for both sexes [27]. To address the inconsistencies in different studies, a recent meta-analysis involving more than 30,000 participants concluded that only MwPD experienced more SD than controls [28]. However, even this finding should be interpreted cautiously due to the heterogeneity in the included studies.

Also, how SH is affected by PD tends to differ according to gender: women reported more difficulties with arousal, reaching an orgasm, low sexual desire, and sexual dissatisfaction, while men reported erectile dysfunction, sexual dissatisfaction, premature ejaculation, and difficulties in reaching an orgasm [14,29]. Libido is more reduced in both sexes [30], but WwPD were more affected than MwPD [21]. The progression of SD may also be affected by gender: male sexual dysfunction increases in 7-year follow-up while no changes in female sexual function were found [31]. Finally, hypersexuality, defined as increased libido and repetitive behaviors toward sexual gratification outside the accepted social and personal bounds, is more prevalent in MwPD (5.2%) than in WwPD (0.5%) [32].

Although some motor symptoms may increase the difficulty in sexual activities [33], the relationship between disease severity and SD varies by study. While some studies showed that SD increases with disease progression [18,22,30], others showed no correlation between them [16,19,34,35]. The disease stages and the treatment time were correlated with significant changes in libido and sexual activity, especially in WwPD [21]. Reduced motor disability level is associated with higher sexual activity in MwPD [9] and sexual satisfaction in MwPD but not in WwPD [36]. Furthermore, some studies found a significant association between disease severity and SD [21,22,24,37], but others did not [13,16,17,30,26].

Some studies found no correlation between depression and SD [19,35,38] or cessation of sexual activity [14], while others point that depression is the only factor associated with sexual and marital satisfaction [8] and a unique common factor to explain sexual dissatisfaction in PwPD [36]. Depression severity was the main predictor factor for loss of libido in PwPD [24]. A 2-year longitudinal prospective study [9] found that lower depressive level was associated with higher sexual activity in MwPD, but not in WwPD. Depression was also correlated with patient’s and partner’s relationship dissatisfaction in EOPD [39].

The impact of cognitive decline and SH has been little investigated. Kummer et al. found that cognitive decline, although associated with libido loss in MwPD, had no predictive value [24]. Picillo et al. [9] found no correlation between cognition capacity and sexual activity in PwPD. Other NMS, such as fatigue, lack of motivation, anxiety, apathy, and sadness, were associated with SD, mainly in WwPD [35,40].

The association between autonomic dysfunction and dopamine repositioning with SH is also controversial. Despite the high prevalence of autonomic dysfunction, this, and the daily dosage levodopa (DDL) was unrelated to overall sexual function [26]. In contrast, other studies demonstrated that autonomic dysfunction was correlated with SD [41], particularly urinary dysfunction [20], and a higher dose of L-dopa was associated with decreased desire in WwPD [14].

Among the social factors, several studies have shown that SD affects both the patient and the sexual partner [8,29]. The couple relationship quality (CRQ) has been bidirectionally related to SH. Brown et al. [8] showed that couples where one of the partners had PD who reported a better marital relationship were more satisfied with their sex life and vice versa. However, satisfaction with marital relationships varied between genders: although MwPD rejected sex less often and reported stronger desire [42], they showed more dissatisfaction with their sex life and marital relationship compared with WwPD [14].

To our knowledge, only two studies focused exclusively on SH in WwPD. Welsh et al. [34] found that WwPD reported more dissatisfaction with the quality of their sexual experiences and were less satisfied with their sexual relationships and with their partners than control. More recently, Varanda et al. [16] found higher prevalence of SD in WwPD than controls, and while age and depression severity were predictors of SD, disease duration, disability level, disease evolution, and antiparkinsonian medication were not. In a recent study, Bronner et al. [43] recommended that more studies focus on WwPD since they experience from the first PD stages and frequently refrain from sharing their sexual concerns.

Given the extensive range of factors that may affect SH and the low number of studies that have focused on the impact of different factors on SH in WwPD, this study aimed to investigate motor, non-motor, and social aspects in the sexual health of WwPD.

## Materials and Methods

### Study Design and Participants

A cross-sectional study including 100 women with PD. The eligibility criteria were (a) women; (b) confirmed diagnosis of idiopathic PD according to the UK Parkinson’s Disease Society Brain Bank diagnostic criteria [44], age above 21 years; (c) reporting active sexual function in the last six months; and (d) have access to telephone or internet and agree to participate in the study. The non-eligibility criteria were (a) presence of neurological disorders other than PD; and (b) presence of dementia, speech, and hearing disorders that could impair the remote interview.

### Recruitment

Participants were recruited from the contacts of the AMPARO network (www.amparo.numec.prp.usp.br). Initial eligibility was identified through calls and/or telephone messages. Subsequently, information about the study procedures was passed on, and they were invited to consent to participate.

This study was approved by the Ethics Committee of the Federal University of Amapa, Macapa, Brazil (#CAAE 39971420.0.0000.0003) and conducted in accordance with the Helsinki Declaration.

### Study Procedures

Interviews were conducted using a structured form and validated instruments. Participants were asked to indicate the best day and time for the remote interview. To avoid tiredness of participants, the interviews were conducted in two stages, each lasting approximately 30-35 minutes, and performed by the same person. The interval between interviews was less than 7 days.

The study flow is demonstrated in Figure 1.

**FIG. 1.** The schematic study designs. T-MoCA, Telephone-Montreal-Cognitive-Assessment; MDS-UPDRS, Movement Disorder Society – Unified Parkinson’s Disease Rating Scale; FSS, Fatigue Severity Scale; RSES, Rosenberg’s Self-esteem Scale; PDSS, Parkinson’s Disease Sleep Scale; DAS, Dyadic Adjustment Scale; BDI, Beck Depression Inventory; FSFI, Female Sexual Function Index; SQ-F, Sexual Quotient – Female.

### Instruments

#### Telephone - Montreal Cognitive Assessment (T-MoCA)

The T-MoCA is an adapted version of the MoCA 30 administered by phone. It contains only the items that do not require the use of pencil and paper or visual stimuli, so its maximum score is 22 [45].

#### Movement Disorder Society - Unified Parkinson’s Disease Rating Scale (MDS-UPDRS)

The MDS-UPDRS is a tool to measure PD severity and progression based on the difficulties presented in the last seven days. In this study, only Part I and Part II were used [46].

#### Fatigue Severity Scale (FSS)

The FSS is a non-specific rating scale often used in PD. This instrument contains 9 questions with answers ranging from 1 (strongly disagree) to 7 (strongly agree). The total score is the mean of the 9 questions, and higher scores indicate higher fatigue degrees [47].

#### Rosenberg’s Self-esteem Scale (RSES)

The RSES comprises 10 questions to assess self-esteem. Questions are answered on a Likert-type scale of how many points range from strongly agree, agree, disagree, and strongly disagree [48].

#### Parkinson’s Disease Sleep Scale (PDSS)

The PDSS is a specific scale for assessing sleep disorders in PD [49]. It comprises 15 questions associated with sleep disorders based on the last week. The score ranges from always (0) to never (10), except for question 1, whose scale ranges from poor (0) to excellent (10). The maximum score is 150 [50].

#### Dyadic Adjustment Scale (DAS)

The DAS is a self-report tool to assess marital adjustment. It consists of 4 subscales of dyadic satisfaction, cohesion, consensus, and affectional expression, with scores ranging from 0 to 5. The total score (sum of all questions) ranges from 0 to 151. Higher scores indicate a better relationship. A total score below 101 indicates dissatisfaction [51].

#### Beck Depression Inventory (BDI)

The BDI is widely used to screen for depression and to measure behavioral manifestations and severity of depression. The BDI comprises 21 questions about quality and depressive symptoms, ranging from 0-63 points (0-13 no depression, 14-19 mild depression, 20-28 moderate, 29-63 severe depression). The total score is the sum of all values. The BDI has been described as a valid instrument to assess depressive symptoms in PD [52].

#### Female Sexual Function Index (FSFI)

The FSFI is a multidimensional self-report instrument that assesses women sexual function in the last four weeks [53]. It comprises 6 domains: desire, arousal, lubrification, orgasm, satisfaction, and pain. The 19 items of the FSFI use a 5-point Likert scale ranging from 1 to 5, with higher scores indicating greater levels of sexual functioning on the individual item. Desired domain is the only domain that can be used independently: scores equal or below 5 indicate the diagnostic criteria for Hypoactive Sexual Desire Disorder (HSDD) [54], and a score inferior to 26.5 indicates SD.

#### Sexual Quotient – Female (SQ-F)

The Sexual Quotient – Female (SQ-F) is an instrument designed to assess emotional and functional aspects related to the sexual performance/satisfaction of women in the last 6 months. Through 10 self-report questions, it evaluates sexual desire and interest/desire, preliminary, excitement and harmony with the partner, comfort, and orgasm and satisfaction. Scores for each question range from 0-5 with higher values indicating better sexual performance/satisfaction. Scores equal or above 60 indicate SD[55].

#### Statistical Analyses

Descriptive statistical analysis was used for demographic and clinical data. The Spearman Rank Order Correlation was used to test correlations among age, socio-economic classification, DDL, disease duration, disease onset, H&Y, T-MoCA, MDS-UPDRS I, MDS-UPDRS II, FSS, RSES, PDSS, DAS, BDI with FSFI and SQ-F and between FSFI and SQ-F.

Additionally, a multiple regression model as predictor variables included all factors that reached a significant statistical correlation with FSFI and SQ-F (response variable). Differences were considered statistically significant for *p* < 0.05. The statistical analyses were performed using Statistica Version 13 (TIBCO Software Inc. USA).

## Results

The participants’ demographic and clinical characteristics are shown in Table 1.

**Table 1:** Demographic and clinical features of the participants.

| **Variables** | **Mean** | **Std. Dev.** |
| --- | --- | --- |
| Age | 53.45 | 8.3 |
| Socio-economic condition | 35.11 | 10.5 |
| DDL | 593.18 | 288.3 |
| Disease duration | 6.31 | 5.1 |
| H&Y | 1.70 | 0.7 |
| T-MoCA | 18.29 | 2.1 |
| MDS-UPDRS I | 11.90 | 7.7 |
| MDS-UPDRS II | 11.79 | 7.4 |
| FSS | 37.35 | 16.9 |
| RSES | 7.11 | 5.7 |
| PDSS | 98.47 | 25.2 |
| DAS | 103.65 | 20.4 |
| BDI | 11.89 | 7.9 |

DDL, Daily dosage levodopa; H&Y, Hoehn and Yahr Stage; T-MoCA, Telephone-Montreal-Cognitive-Assessment; MDS-UPDRS, Movement Disorder Society – Unified Parkinson’s Disease Rating Scale; FSS, Fatigue Severity Scale; RSES, Rosenberg’s Self-esteem Scale; PDSS, Parkinson’s Disease Sleep Scale; DAS, Dyadic Adjustment Scale; BDI, Beck Depression Inventory.

Additionally, 45% of participants were in stage 1, 40% in stage 2, and 15% in stage 3 according to H&Y classification; 36% of participants presented EOPD, and 67% were in a postmenopausal lifetime.

### Short-term sexual health (STSH)

The mean score in FSFI was 20.93±4.9, below the minimal score adopted to identify sexual dysfunction (cut-off score = 26,5). Furthermore, the mean score in the Desire domain was 3.63, a value below the cut-off score for HSDD diagnosis. The mean scores by domain can be seen in Table 2.

**Table 2:** FSFI domain scores.

| **Domain** | **Question/factor** | **Score** | **Mean/Std.Dev.** |
| --- | --- | --- | --- |
| Desire | 1,2 (0,6) | 1.2 – 6 | 3.63±1.1 |
| Arousal | 3,4,5,6 (0,3) | 0 – 6 | 3.11±1.4 |
| Lubrication | 7,8,9,10 (0,3) | 0 – 6 | 3.63±0.9 |
| Orgasm | 11,12,13 (0,4) | 0 – 6 | 3.43±1.0 |
| Satisfaction | 14,15,16 (0,4) * | 0.8 – 6 | 2.51±1.2 |
| Pain | 17,18,19 (0,4) | 0 – 6 | 4.59±2.1 |
| Total scores |  | 2 – 36 | 20.93±4.9 |

\*Variation item 14 = 0 – 5; item 15 and 16 = 1 – 5. FSFI, Female Sexual Function Index.

The total scores obtained by FSFI had a weak positive statistically significant correlation with RSES and BDI scores and a weak negative statistically significant correlation with DAS scores (Table 3).

**Table 3:** Correlation between FSFI, demographic and clinical variables.

| **Variables** | **R** | **p** |
| --- | --- | --- |
| Age | R =.037 | p =.757 |
| Socio-economic condition | R =-.054 | p =.651 |
| Disease onset | R=.049 | p=.623 |
| Disease duration | R =.005 | p =.965 |
| H&Y | R =.091 | p =.443 |
| DDL | R =.151 | p =.205 |
| Postmenopausal lifetime | R=.030 | p=.762 |
| TMoCA | R =-.214 | p =.070 |
| MDS-UPDRS I | R =.211 | p =.074 |
| MDS-UPDRS II | R =.063 | p =.599 |
| FSS | R =.115 | p =.332 |
| PDSS | R =-.069 | p =.563 |
| **DAS** | **R =-.236** | **p =.046** |
| **BDI** | **R =.324** | **p =.005** |
| **RSES** | **R =.150** | **p =.135** |

FSFI, Female Sexual Function Index; H&Y, Hoehn and Yahr Stage; DDL, Daily dosage levodopa; T-MoCA, Telephone-Montreal-Cognitive-Assessment; MDS-UPDRS, Movement Disorder Society – Unified Parkinson’s Disease Rating Scale; FSS, Fatigue Severity Scale; PDSS, Parkinson’s Disease Sleep Scale; DAS, Dyadic Adjustment Scale; BDI, Beck Depression Inventory; RSES, Rosenberg’s Self-esteem Scale.

### Long-term sexual health (LTSH)

The mean score in SQ-F was 72.92±21.2, above the score adopted to identify sexual dysfunction (cut-off score = 60). The mean scores by domain can be seen in Table 4.

**Table 4:**
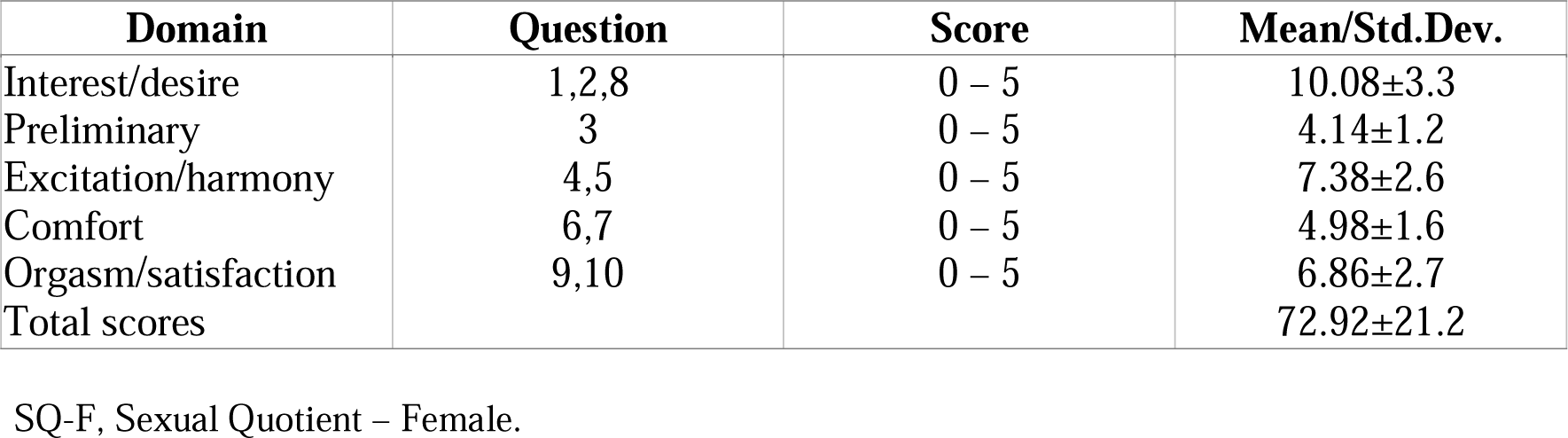
SQ-F domain scores.

| **Domain** | **Question** | **Score** | **Mean/Std.Dev.** |
| --- | --- | --- | --- |
| Interest/desire | 1,2,8 | 0 – 5 | 10.08±3.3 |
| Preliminary | 3 | 0 – 5 | 4.14±1.2 |
| Excitation/harmony | 4,5 | 0 – 5 | 7.38±2.6 |
| Comfort | 6,7 | 0 – 5 | 4.98±1.6 |
| Orgasm/satisfaction | 9,10 | 0 – 5 | 6.86±2.7 |
| Total scores |  |  | 72.92±21.2 |

SQ-F, Sexual Quotient – Female.

The total score obtained by SQ-F had a weak positive statistically significant correlation with T-MoCA and PDSS, a weak negative statistically significant correlation with T-MoCA, MDS-UPDRS I, a strong positive statistically significant correlation with DAS, and a strong negative statistically significant correlation with BDI (Table 5).

**Table 5:** Correlation between SQ-F, demographic and clinical variables.

| **Variables** | **R** | **p** |
| --- | --- | --- |
| Age | R = -.092 | p =.442 |
| Race | R = -.009 | p =.937 |
| Disease duration | R = -.222 | p =.060 |
| Disease onset | R= -.029 | p=.769 |
| H&Y | R=.096 | p=.340 |
| DDL | R = -.216 | p =.067 |
| Postmenopausal lifetime | R=. -.008 | p=.936 |
| **T-MoCA** | **R = .311** | **p =.008** |
| **MDS-UPDRS I** | **R = -.299** | **p =.011** |
| MDS-UPDRS II | R = -.152 | p =.201 |
| FSS | R = -.220 | p =.063 |
| **PDSS** | **R =.363** | **p =.002** |
| **RSES** | **R = -.273** | **p =.020** |
| **DAS** | **R =.520** | **p =.000** |
| **BDI** | **R = -.479** | **p =.000** |

H&Y, Hoehn and Yahr Stage; Daily dosage levodopa; T-MoCA, Telephone-Montreal-Cognitive-Assessment; MDS-UPDRS, Movement Disorder Society – Unified Parkinson’s Disease Rating Scale; FSS, Fatigue Severity Scale; PDSS, Parkinson’s Disease Sleep Scale; RSES, Rosenberg’s Self-esteem Scale; DAS, Dyadic Adjustment Scale; BDI, Beck Depression Inventory.

The variables DAS, BDI, and RSES were included in the multiple regression model, with only the BDI remaining as a predictor variable for the FSFI, which presented an R^2^=.10 and Beta =.15.

Furthermore, the multiple regression model included the variables T-MoCA, MDS-UPDRS I, PDSS, DAS, BDI, and RSES. T-MoCA, and DAS scores remained in the final model as independent predictors with R^2^=.36, and Beta= 2.67 and .47, respectively.

Finally, a moderate negative correlation was found between FSFI and SQ-F (r=.46; p<.0001).

## Discussion

Although SH is a fundamental aspect of the overall health and well-being of individuals, families, and countries [56], few studies have investigated SH in PwPD, particularly women. To the best of our knowledge, the present study was the first aiming to investigate the impact of the motor, non-motor, and social aspects on the short and long-term SH of WwPD by different but complementary instruments developed to assess sexual function.

Our results showed that, among all investigated factors, mental aspects, mainly depression and cognition, and social aspects as CRQ significantly impacted SH of WwPD. Surprisingly, age, disease onset, and disease severity measured by H&Y, disease duration, and DDL had no significant impact on SH. In addition, NMS severity and motor disability level measured by parts I and II of MDS-UPDRS respectively, also had no impact on SH.

Taken together, several considerations can be made based on the results.

In the present study, besides using a female-specific instrument (FSFI) that assess SH in the last four weeks, our findings are further strengthened by using the SQ-F, providing a complementary qualitative approach to female sexual function based on the last six months. Although the results of both instruments showed a moderate correlation between them and with depression, self-esteem, and CRQ, the SQ-F was correlated with a larger number of factors than FSFI, including cognition and severity of NMS. Furthermore, the results from multiple regression models confirmed that FSFI and SQ-F scores could be predicted by different factors: while depression severity was the unique able to predict the FSFI, the global cognitive capacity and that CRQ were the best predictors for the SQ-F. Based on this evidence, although most previous studies investigating SH in WwPD had used FSFI [12,16,20,27,30,36,43], the addition of an evaluation over a longer term by SQ-F may offer a more comprehensive disease impact on SH..

According to our findings, 88% of WwPD suffered from SD based on FSFI and 78% based on SQ-F and, according to the Desire domain of the FSFI, 88% of women meet the diagnostic criteria for HSDD. Among the studies investigating the SD in WwPD, Varanda et al. [16] found SD in 86,9% of WwPD and 79,0% in controls. Kotková and Weiss [36] found 75,8% of WwPD reported SD. Furthermore, Pedro et al. [12] showed that SD was reported in 77,8% of WwPD treated with DBS-STN (Deep brain stimulation – subthalamic nucleus). Finally, Raciti et al [30] found that only 53% of WwPD reported SD.

Most importantly, the observed absence of a correlation between SD and age or disease severity suggests that this kind of NMS exists even in young WwPD. Several studies showed SD and sexual satisfaction in PwPD worsened with aging and disease progression [13,14,15,16,34,35]. In the present study, 36% of women were younger than 50 years, attending EOPD criteria. However, there was no correlation between disease onset and the STSH and LTSH. Previous studies have shown that SD is a frequent NMS in EOPD and may be underestimated [18,20]. Despite the increased prevalence of SD in postmenopausal health women [57], we found no correlation between postmenopausal and SD, indicating that SH is an essential issue for WwPD regardless of age, disease onset, and disease stages. Thus, a proper therapeutic intervention should be available from the disease beginning until the moderate PD stages.

Dopaminergic medication has been correlated with SD in MwPD [5] and WwPD [14]. However, in the present study, the DDL was not associated with SH. Varanda et al. [16] also found no difference in DDL and the presence or absence of SD in women with PD.

Depression severity was the unique factor correlated with STSH and LTSH, although able to predict only short-term sexual dysfunction. Previous studies that have investigated the relationship between SD and depression in PD showed that depression severity was associated with loss of libido sexual [24], sexual and marital satisfaction [8,36], decreased intimacy in women. The results from the two previous studies, which included only WwPD, confirm the negative impact of depression on SH. Welsh [34] showed that WwPD presented higher depression than controls, associated with decreased libido. Varanda et al. [16] showed that depression severity could predict SH in WwPD. Given that 54.9% of WwPD report depressive symptoms and 15.7% of them have major depressive symptoms [58], deepening the investigation of the relationship between depression and SD in PwPD is a critical issue to be investigated in further studies.

While depression was an independent factor in predicting the STSD, better global cognitive capacity and quality of couple relationship were the factors able to predict the LTSH. Although cognitive decline is one of the most prevalent NMS in PD [59], with about 20% to 30% of PwPD already showing mild cognitive impairment at diagnosis time [60,61], studies investigating the relationship between cognitive decline and SH in PD are scarce. Hand et al. found no association between SD and cognition [19], while Kummer found no predictive value of cognition for libido loss, although both were correlated [24]. In both studies, cognition was evaluated by the Mini-Mental State Examination (MMSE). The MoCA has been considered a more proper tool for assessing to PD’s global cognitive capacity than MMSE. Thus, better MoCA sensibility for mild cognitive decline [62], may explain, at least in part, the present results. The slow decline in the cognitive capacity observed in PD may affect the LTSD but not the STSD. Supposedly, intervention to decrease cognitive decline may prevent the future decrease in SD. This issue should be investigated in further studies.

In the present study, DAS scores were correlated with short and LTSH, and better CRQ predicted better LTSH. A few studies investigated the relationship between CRQ and SH. Brown et al. showed that most WwPD and their partners had scores within the normal range for marital satisfaction. In contrast, the MwPD and their partners found the most significant marital dissatisfaction and that higher marital satisfaction was associated with better SD for both women and men [8]. Wielinski et al. have also shown that sexual dissatisfaction is correlated with relationship dissatisfaction in PwPD [39]. These findings reinforce the importance of offering intervention for the couple, instead of for the patient only, to improve or keep the CRQ even facing chronic and disabling disease.

Some previous studies have found associations between other NMS and SD [40]. However, the MDS-UPDRS I score, although correlated with LTSH, had no predictive value in the present study. The same was observed for self-esteem: despite some features like drooling, excessive sweating, seborrhea, and hypomimia may decrease self-esteem [5], the RSES scores, although correlated with short and LTSH, had no predictive value for both.

The present study presents some strengths: the use of multidimensional generic instruments to assess STSH and LTSH, CRQ, depression, and cognition as disease-specific instruments that comprehensively assess motor and non-motor dysfunction in PD.

Our study has some limitations. First, the study’s cross-sectional design may represent the principal limitation because of information based on data gathered for a specific point in time. However, including different tools to assess the STSH and LTSH may have minimized this limitation. In addition, the sample could not be large enough to allow a large generalization to the PD population. However, including participants from 5 different geographical areas, with sizable sociocultural differences, of a country with a continental area may have minimized this limitation. Finally, the absence of women in advanced stages with PD who reported active sexual life may also be considered a study limitation.

## Conclusion

SH was impaired in most WwPD, regardless of age, disease onset or disease severity. Depression severity predicted the STSD, while global cognitive capacity and CRQ predicted the LTSD. Therefore, an interdisciplinary approach to improve depression, cognition, and CRQ and address complex real-life issues should be available for WwPD since the early disease stages to avoid the decline in SH.

## Data Availability

All data produced in the present work are contained in the manuscript.

http://redeamparo.com.br

## Acknowledgments

This article was produced as part of the activities of the FAPESP Research, Innovation, and Dissemination Center for Neuromathematics (grant #<u>2013/ 07699-0</u>, S. Paulo Research Foundation). This article was supported in part by the Coordenação de Aperfeiçoamento de Pessoal de Nível Superior, CAPES, Brazil.

## Data Availability Statement

The data included in this study are available on reasonable request to the corresponding author. The data are not publicly available due to the inclusion of information that could compromise the participants’ privacy.

## Author Roles

(1) Research Project: A. Conception, B. Organization, C. Data collection; (2) Manuscript Preparation: A. Writing of the First Draft, B. Review and Critique.

K.C.C.N.: 1A, 1B, 1C, 2A, 2B

I.A.P.S.N.: 1B, 2A, 2B

B.R.A.S.: 1A, 1B, 1C R.A.G.: 1C

T.S.M.: 1C

G.F.S.: 1C

B.E.A.S.: 1C

A.F.H.: 2B

A.C.R.: 2B

R.S.: 2B

M.E.P.P.: 1A, 1B, 2A, 2B

## Disclosures

**Ethical Compliance Statement:** Participants gave informed consent, and the study was approved by the Ethics Committee of the Federal University of Amapa, Macapa, Brazil (#CAAE 39971420.0.0000.0003) and conducted in accordance with the Helsinki Declaration. We confirm that we have read the Journal’s position on issues involved in ethical publication and affirm that this work is consistent with those guidelines.

**Funding Sources and Conflict of Interest:** This article was produced as part of the activities of FAPESP Research, Innovation, and Dissemination Center for Neuromathematics (grant #<u>2013/ 07699-0</u>, S.Paulo Research Foundation). The authors report no conflicts of interest.

**Financial Disclosures from previous 12 months:** None.

## Notes

### Competing Interest Statement

The authors have declared no competing interest.

### Funding Statement

This study did not receive any funding.

